# Plasma CXCL8 and MCP-1 as biomarkers of latent tuberculosis infection

**DOI:** 10.1101/2023.08.07.23293767

**Authors:** Sivaprakasam T Selvavinayagam, Bijulal Aswathy, Yean K Yong, Asha Frederick, Lakshmi Murali, Vasudevan Kalaivani, Karishma S Jith, Manivannan Rajeshkumar, Adukkadukkam Anusree, Meganathan Kannan, Natarajan Gopalan, Ramachandran Vignesh, Amudhan Murugesan, Hong Yien Tan, Ying Zhang, Samudi Chandramathi, Munusamy Ponnan Sivasankaran, Sakthivel Govindaraj, Siddappa N Byrareddy, Vijayakumar Velu, Marie Larsson, Esaki M Shankar, Sivadoss Raju

## Abstract

**Background:** Early detection of latent tuberculosis infection (LTBI) is critical to TB elimination in the current WHO vision of *End Tuberculosis Strategy*.

**Methods:** We investigated whether detecting plasma cytokines could aid in diagnosing LTBI across household contacts (HHCs) positive for IGRA, HHCs negative for IGRA, and healthy controls. We also measured the plasma cytokines using a commercial *Bio-Plex Pro Human Cytokine 17-plex* assay.

**Results:** Increased plasma CXCL8 and decreased MCP-1, TNF-α, and IFN-γ were associated with LTBI. Regression analysis showed that a combination of CXCL8 and MCP-1 increased the risk of LTBI among HHCs to 14-fold.

**Conclusions:** We postulated that CXCL8 and MCP-1 could be the surrogate biomarkers of LTBI, especially in resource-limited settings.

## 1 INTRODUCTION

Tuberculosis (TB) is one of the most devastating infectious diseases, resulting in ∼1.6 million deaths in 2021. Reports suggest that one-fourth of the global population was infected with *Mycobacterium tuberculosis* (*M. tuberculosis* or MTB) in 2021.^1^ Latent TB infection (LTBI) results in persistent immune responses to MTB antigens without any gross evidence of clinical TB.^1, 2^ Estimates suggest that 5-10% of individuals with underlying LTBI might progress to develop active TB disease.^1,3^ According to the World Health Organization’s (WHO) *End Tuberculosis Strategy*, the early detection of LTBI, especially in endemic areas, is key to global TB elimination.^4, 5^ Hence, an improved method to detect LTBI is urgently required,^4,6^ especially in resource-limited settings.

TB diagnostics suffers from lack of a gold standard test.^7^ Until the advent of the interferon-gamma-release assay (IGRA), the tuberculin skin test (TST) was the only tool available.^8,9^ TST uses a purified protein derivative (PPD) antigen.^9^ It endures poor sensitivity for use in immune-compromised individuals and poor specificity due to several confounding factors.^10,11^ IGRA appears more specific than the TST for detecting LTBI.^5,12^ In addition, an individual’s immune status could influence IGRA results as suggested by others.^8,13^ However, IGRA and TST cannot distinguish between active TB and LTBI.^2,11,14^ Hence, to address these limitations, identifying an alternative molecular biomarker is necessary.^5,7^

During MTB infection, macrophages regulate cytokines secreted by T cells.^15^ In concert with complex mycobacterial antigens, cytokines mount protective and pathogenic responses.^16^ Previous literature has suggested that specific cytokines could aid in distinguishing between the stages of disease progression.^5,12^ Here, we investigated whether specific cytokines, apart from IFN-γ, have any association with LTBI for potential use as a surrogate biomarker(s) to detect LTBI among household contacts (HHCs) of individuals with active TB disease.

## 2 METHODS

### 2.1 Ethical approval

This study was performed within the ethical standards of the Declaration of Helsinki. The study procedures and/or protocols were reviewed and approved by the Human Ethics Committee of the Directorate of Public Health and Preventive Medicine Ethical Committee (DPH, IEC 15-10-2022), Chennai, India. All participants provided written informed consent to participate in the study. The flow diagram outlining the study plan is provided in *Figure 1*.

**Figure 1.**
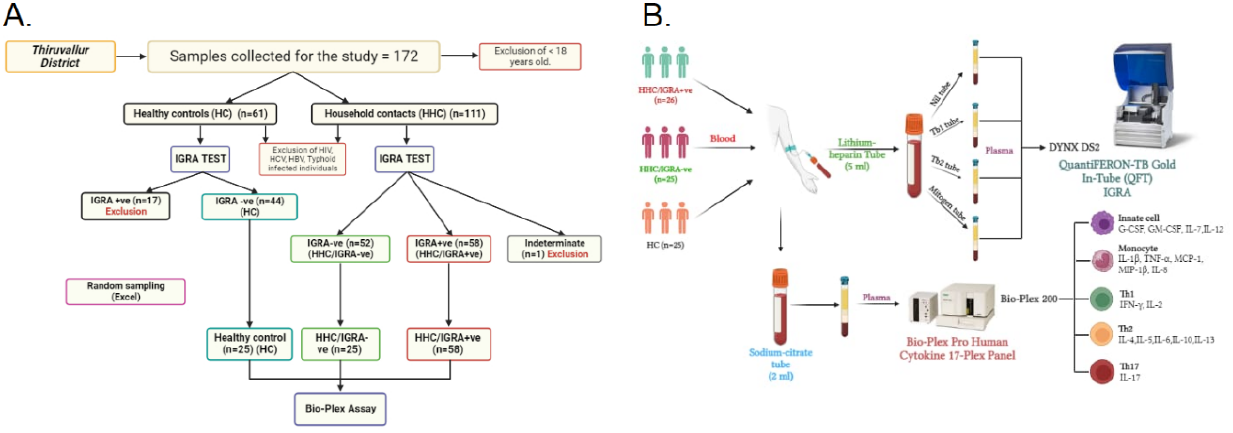
Sampling and study set up. **A)** The flow-chart outlining the cross-sectional study sampling for the LTBI cohort. The study recruited 76 volunteers by a random sampling method, the exclusion criteria’s as well as the IGRA reports (from a total of 172 individuals) who were further divided into three cohorts: Household contacts/IGRA+ve (HHC/IGRA+ve) (n=26 from 58 positive individuals), household contacts/IGRA-ve (HHC/IGRA-ve) (n=25 from 52 negative individuals) and healthy controls (HCs) (n=25 from 44 individuals tested negative for LTBI). HCs were defined as having had no contact with active TB cases and were negative for IGRA. **B)** Blood was drawn in Lithium-heparin tubes and Sodium-citrate tubes. Plasma samples in the Lithium-heparin tubes were subjected to IGRA after transferring them to the QFT tubes. Plasma samples in the Sodium-citrate tubes were subjected to Bio-Plex Luminex Cytokine assay.

### 2.2 Samples and study design

The cross-sectional study recruited 76 volunteers by a random sampling method (from a total of 172 individuals) who were further divided into three cohorts: Household contacts/IGRA+ve (HHC/IGRA+ve) (n=26 from 58 positive individuals), household contacts/IGRA-ve (HHC/IGRA-ve) (n=25 from 52 negative individuals) and healthy controls (HCs) (n=25 from 44 individuals tested negative for LTBI). HCs were defined as having had no contact with active TB cases and were negative for IGRA. Clinico-demographic data was obtained from each participant (*Table 1*).

**Table 1.**
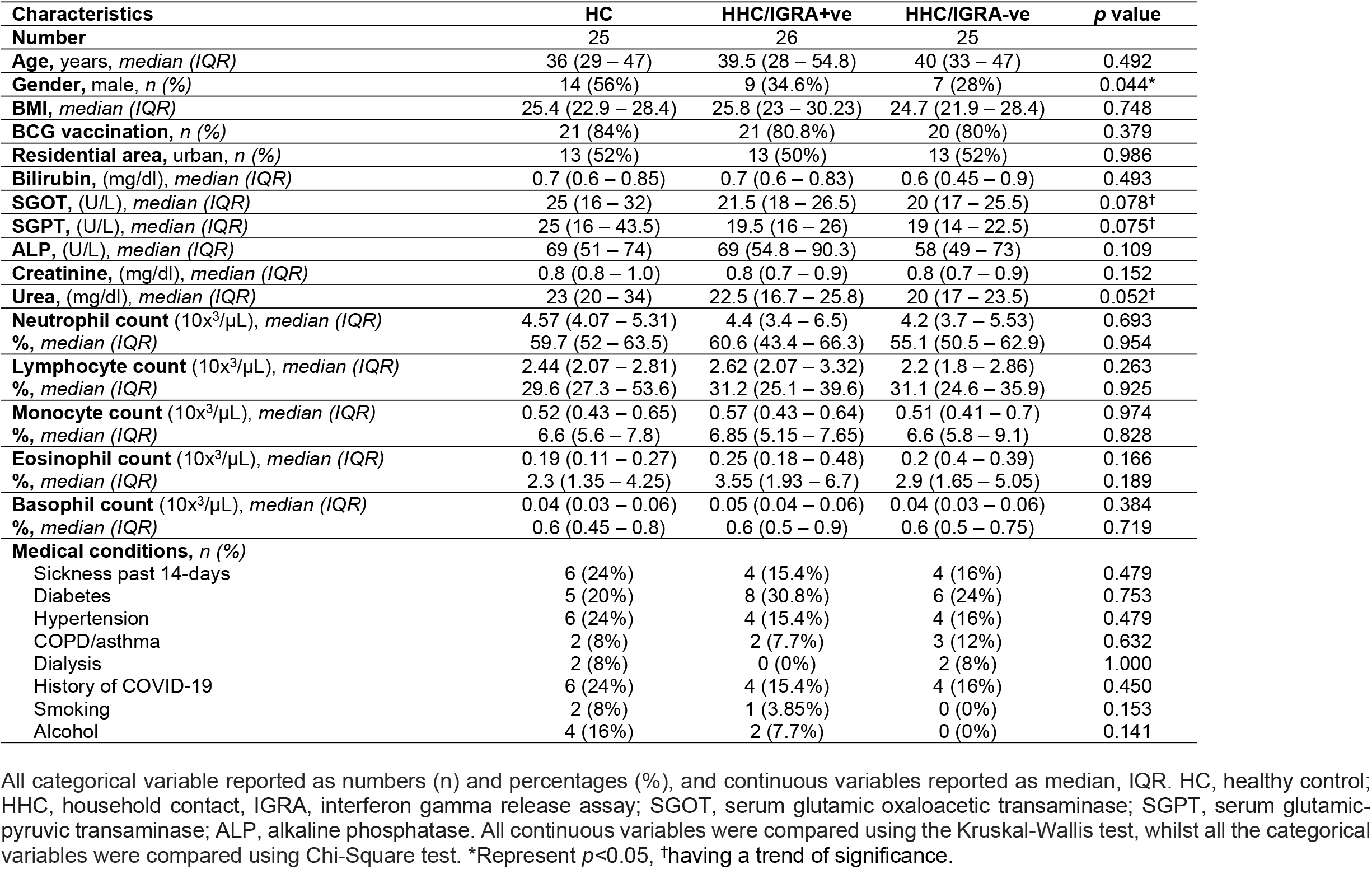
Clinico-demographic characteristics of the study cohort.

| Characteristics | HC | HHC/IGRA+ve | HHC/IGRA-ve | p value |
| --- | --- | --- | --- | --- |
| **Number** | 25 | 26 | 25 |  |
| **Age, years, median (IQR)** | 36 (29 – 47) | 39.5 (28 – 54.8) | 40 (33 – 47) | 0.492 |
| **Gender, male, n (%)** | 14 (56%) | 9 (34.6%) | 7 (28%) | 0.044* |
| **BMI, median (IQR)** | 25.4 (22.9 – 28.4) | 25.8 (23 – 30.23) | 24.7 (21.9 – 28.4) | 0.748 |
| **BCG vaccination, n (%)** | 21 (84%) | 21 (80.8%) | 20 (80%) | 0.379 |
| **Residential area, urban, n (%)** | 13 (52%) | 13 (50%) | 13 (52%) | 0.986 |
| **Bilirubin, (mg/dl), median (IQR)** | 0.7 (0.6 – 0.85) | 0.7 (0.6 – 0.83) | 0.6 (0.45 – 0.9) | 0.493 |
| **SGOT, (U/L), median (IQR)** | 25 (16 – 32) | 21.5 (18 – 26.5) | 20 (17 – 25.5) | 0.078† |
| **SGPT, (U/L), median (IQR)** | 25 (16 – 43.5) | 19.5 (16 – 26) | 19 (14 – 22.5) | 0.075† |
| **ALP, (U/L), median (IQR)** | 69 (51 – 74) | 69 (54.8 – 90.3) | 58 (49 – 73) | 0.109 |
| **Creatinine, (mg/dl), median (IQR)** | 0.8 (0.8 – 1.0) | 0.8 (0.7 – 0.9) | 0.8 (0.7 – 0.9) | 0.152 |
| **Urea, (mg/dl), median (IQR)** | 23 (20 – 34) | 22.5 (16.7 – 25.8) | 20 (17 – 23.5) | 0.052† |
| **Neutrophil count (10x<sup>3</sup>/μL), median (IQR)** | 4.57 (4.07 – 5.31) | 4.4 (3.4 – 6.5) | 4.2 (3.7 – 5.53) | 0.693 |
| **%, median (IQR)** | 59.7 (52 – 63.5) | 60.6 (43.4 – 66.3) | 55.1 (50.5 – 62.9) | 0.954 |
| **Lymphocyte count (10x<sup>3</sup>/μL), median (IQR)** | 2.44 (2.07 – 2.81) | 2.62 (2.07 – 3.32) | 2.2 (1.8 – 2.86) | 0.263 |
| **%, median (IQR)** | 29.6 (27.3 – 53.6) | 31.2 (25.1 – 39.6) | 31.1 (24.6 – 35.9) | 0.925 |
| **Monocyte count (10x<sup>3</sup>/μL), median (IQR)** | 0.52 (0.43 – 0.65) | 0.57 (0.43 – 0.64) | 0.51 (0.41 – 0.7) | 0.974 |
| **%, median (IQR)** | 6.6 (5.6 – 7.8) | 6.85 (5.15 – 7.65) | 6.6 (5.8 – 9.1) | 0.828 |
| **Eosinophil count (10x<sup>3</sup>/μL), median (IQR)** | 0.19 (0.11 – 0.27) | 0.25 (0.18 – 0.48) | 0.2 (0.4 – 0.39) | 0.166 |
| **%, median (IQR)** | 2.3 (1.35 – 4.25) | 3.55 (1.93 – 6.7) | 2.9 (1.65 – 5.05) | 0.189 |
| **Basophil count (10x<sup>3</sup>/μL), median (IQR)** | 0.04 (0.03 – 0.06) | 0.05 (0.04 – 0.06) | 0.04 (0.03 – 0.06) | 0.384 |
| **%, median (IQR)** | 0.6 (0.45 – 0.8) | 0.6 (0.5 – 0.9) | 0.6 (0.5 – 0.75) | 0.719 |
| **Medical conditions, n (%)** |  |  |  |  |
| Sickness past 14-days | 6 (24%) | 4 (15.4%) | 4 (16%) | 0.479 |
| Diabetes | 5 (20%) | 8 (30.8%) | 6 (24%) | 0.753 |
| Hypertension | 6 (24%) | 4 (15.4%) | 4 (16%) | 0.479 |
| COPD/asthma | 2 (8%) | 2 (7.7%) | 3 (12%) | 0.632 |
| Dialysis | 2 (8%) | 0 (0%) | 2 (8%) | 1.000 |
| History of COVID-19 | 6 (24%) | 4 (15.4%) | 4 (16%) | 0.450 |
| Smoking | 2 (8%) | 1 (3.85%) | 0 (0%) | 0.153 |
| Alcohol | 4 (16%) | 2 (7.7%) | 0 (0%) | 0.141 |

All categorical variable reported as numbers (n) and percentages (%), and continuous variables reported as median, IQR. HC, healthy control; HHC, household contact, IGRA, interferon gamma release assay; SGOT, serum glutamic oxaloacetic transaminase; SGPT, serum glutamic-pyruvic transaminase; ALP, alkaline phosphatase. All continuous variables were compared using the Kruskal-Wallis test, whilst all the categorical variables were compared using Chi-Square test. \*Represent $p < 0.05$ , †having a trend of significance.

Blood samples (lithium heparin (5 ml) and sodium citrate (2 ml) BD Vacutainer tubes) were obtained from 172 individuals who were divided primarily into two cohorts: HCs and HHCs. As per the manufacturer’s instructions, all 172 samples were subjected to a commercial QuantiFERON-TB Gold In-Tube Assay (Cat. No.: 622130, Qiagen, USA). Briefly, heparinized plasma was aliquoted into four QFT-Plus blood tubes, viz., nil tube, TB antigen tube 1 (TB1) possessing ESAT-6 and CFP-10 peptides to activate CD4+ T cells, TB antigen tube 2 (TB2) that includes ESAT-6 and CFP-10 peptides to activate CD8+ T cells and mitogen tube as a positive control. The aliquoted tubes were mixed ∼10 times before 16-24 hours of incubation for 15 min at 3000 rpm. Subsequently, the separated plasma was subjected to an in-built ELISA to detect IFN-γ. The results were calculated using QFT Plus analysis software by analyzing the IFN-γ level of the post-reaction supernatant. The results were interpreted as positive, negative, or indeterminate [4, 9].

To quantify the levels of various plasma cytokines, we used a commercial Bio-plex Pro Human Cytokine 17-plex assay (Bio-Rad Laboratories, Hercules, CA) that uses sodium-citrated plasma without TB antigen stimulation. The kit measures the following cytokines: IL-1β, IL-2, IL-3, IL-4, IL-5, IL-6, IL-7, CXCL8, IL-10, IL-12, IL-13, IL-17A, G-CSF, GM-CSF, IFN-γ, MCP-1, MIP-1 and TNF-α) as per the manufacturer’s instructions. The data were analyzed using the Bio-plex Manager Software Ver.6.1.

### 2.3 Statistical analysis

The comparison of the group differences was made using the Mann-Whitney test. The Kruskal-Wallis test was used for testing statistical significance with the median values. The Chi-Square test was used to compare the categorical variables. Receiver operating characteristic (ROC) curves were drawn to define the diagnostic performance of the biomarker. The best cut-off value (sum of sensitivity and specificity divided by 100) was chosen to maximize Youden’s index and the AUC. GraphPad Prism 6.0 (GraphPad Software, San Diego, CA), SPSS version 20.0, and Microsoft Excel 2019 were used for the statistical evaluation. The level of significance was *p*<0.05.

## 3 RESULTS

### 3.1 Cohort characteristics

The group characteristics revealed considerable differences between the sexes, particularly among the study’s male population. SGOT, SGPT, and urea levels showed a trend of significant difference among the groups. Our study had 13 individuals in the HHC/IGRA+ve group (n=26) with underlying medical conditions. The socioeconomic characteristics of the cohort are presented in *Figure 2*.

**Figure 2.**
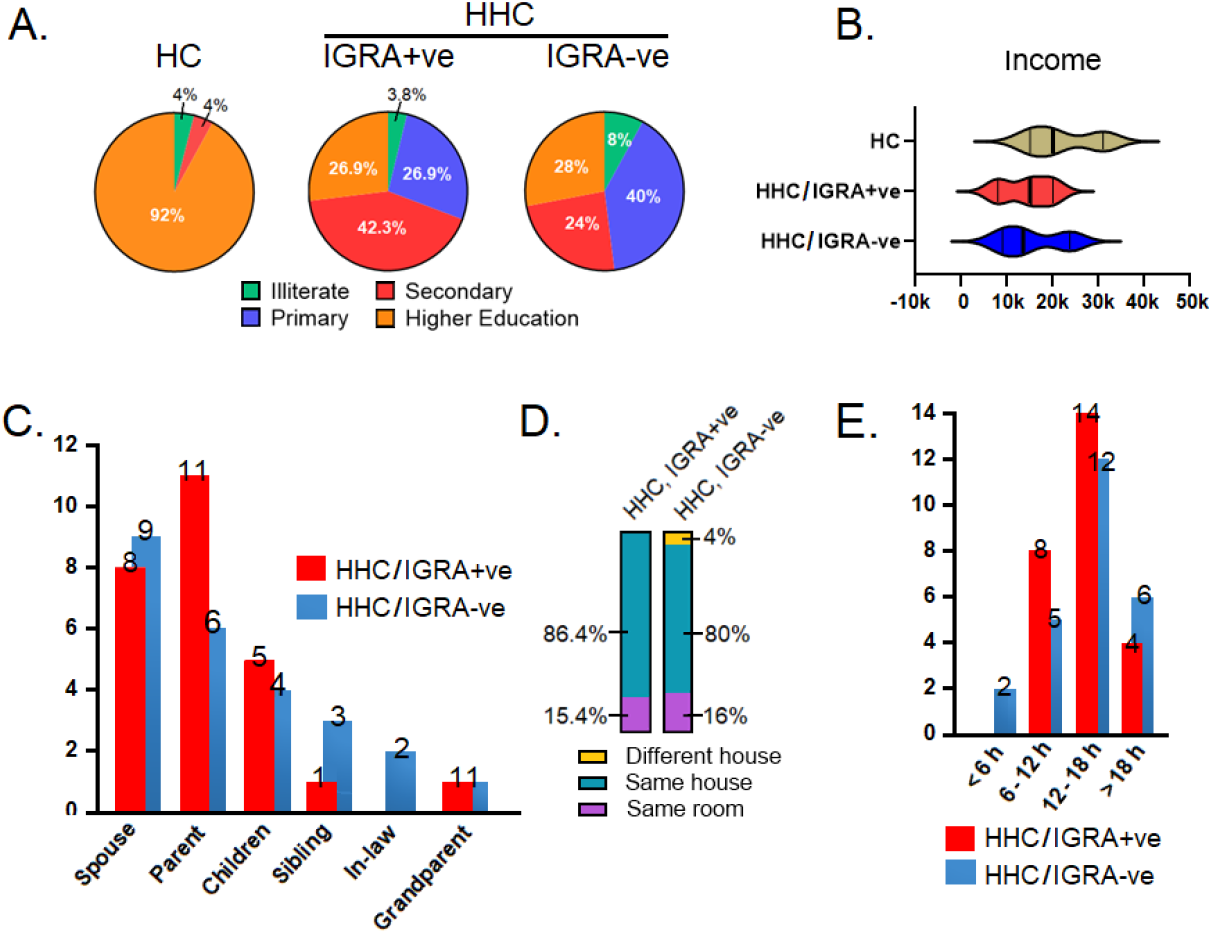
Socioeconomic characteristics of cohort participants. **A)** Education level of participants. **B)** Family annual income of the participants. **C)** Relationship of the active TB index case with the participants. **D)** Proximity of the participants with the active TB index case. **E)** Contact duration of the participants with the active TB index case. HC, healthy control; HHC, household contact; IGRA, interferon gamma releasing assay

### 3.2 An increase of CXCL8 and a decrease of MCP-1, TNF-α, and IFN-γ were associated with LTBI

Seven of the 17 cytokines measured using the Bio-Plex Luminex cytokine assay remained undetectable, viz., IL-2, IL-4, IL-5, IL-7, IL-12, G-CSF, and GM-CSF. Hence, we incorporated only the remaining ten cytokines in the analysis. Of the monocyte-derived cytokines, CXCL8, MCP-1, and TNF-α showed significance. MIP-1 and IL-1 did not show any significant difference. Similarly, IFN-γ showed a significant difference among the T-cell-derived cytokines. IL-6, IL-10, IL-17A, and IL-13 did not reveal any marked differences. CXCL8 levels in HHC/IGRA+ve and HHC/IGRA-ve were higher than HCs (*p*<0.05). MCP-1 levels in HHC/IGRA+ve and HHC/IGRA-ve were lower than the HCs (*p*<0.01). TNF-α levels in the HCC/IGRA+ve and HHC/IGRA-ve were lower than HCs (*p*<0.001). IFN-γ concentrations in HHC/IGRA+ve and HCC/IGRA-ve were lower than HCs (*Figure 3*).

**Figure 3.**
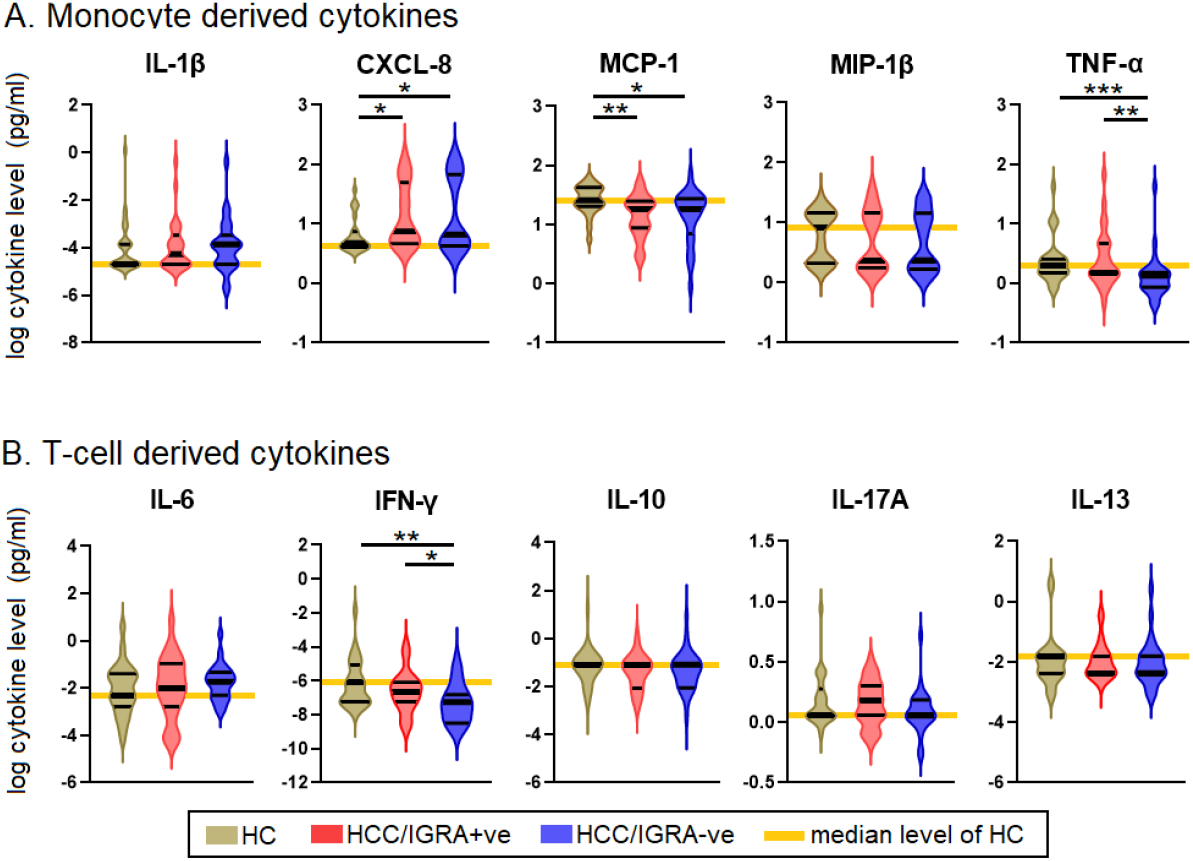
Comparison of the levels of cytokines among HC, HHC/IGRA+ve and HHC/IGRA-ve individuals. **A)** Monocyte-derived cytokines **B)** T cell-derived cytokines. IL, interleukin; MCP-1, monocyte chemoattractant protein-1; MIP-1β, macrophage inflammatory protein-1 beta; TNF-α, tumor necrosis factor alpha; IFN-γ, interferon gamma. *, ** and *** represent *p<*0.05, <0.01, <0.001, respectively.

### 3.3 CXCL8 and MCP-1 predicted the risk of the development of LTBI

To assess the suitability of CXCL8, MCP-1, TNF-α, and IFN-γ as surrogate biomarkers to detect LTBI, receiver-operating characteristics (ROC) analysis was performed between HHC/IGRA+ve and HCs. Our analysis showed that CXCL8 and MCP-1 could predict LTBI with the area under the curve (AUC) of 0.6885; *p*=0.0210 and 0.7392; *p*=0.0034, respectively. However, TNF-α and IFN-γ did not predict LTBI (data not shown). From the ROC analysis, we determined the cut-off for CXCL8 and MCP-1. The cut-off value for CXCL8 was >6.2 pg/ml, while MCP-1 was <22.56 pg/ml. Though the AUC for CXCL8 and MCP-1 were seemingly low when we combined both CXCL8 and MCP-1, the AUC was 0.9393; *p*<0.0001 (*Figure 4A*).

**Figure 4.**
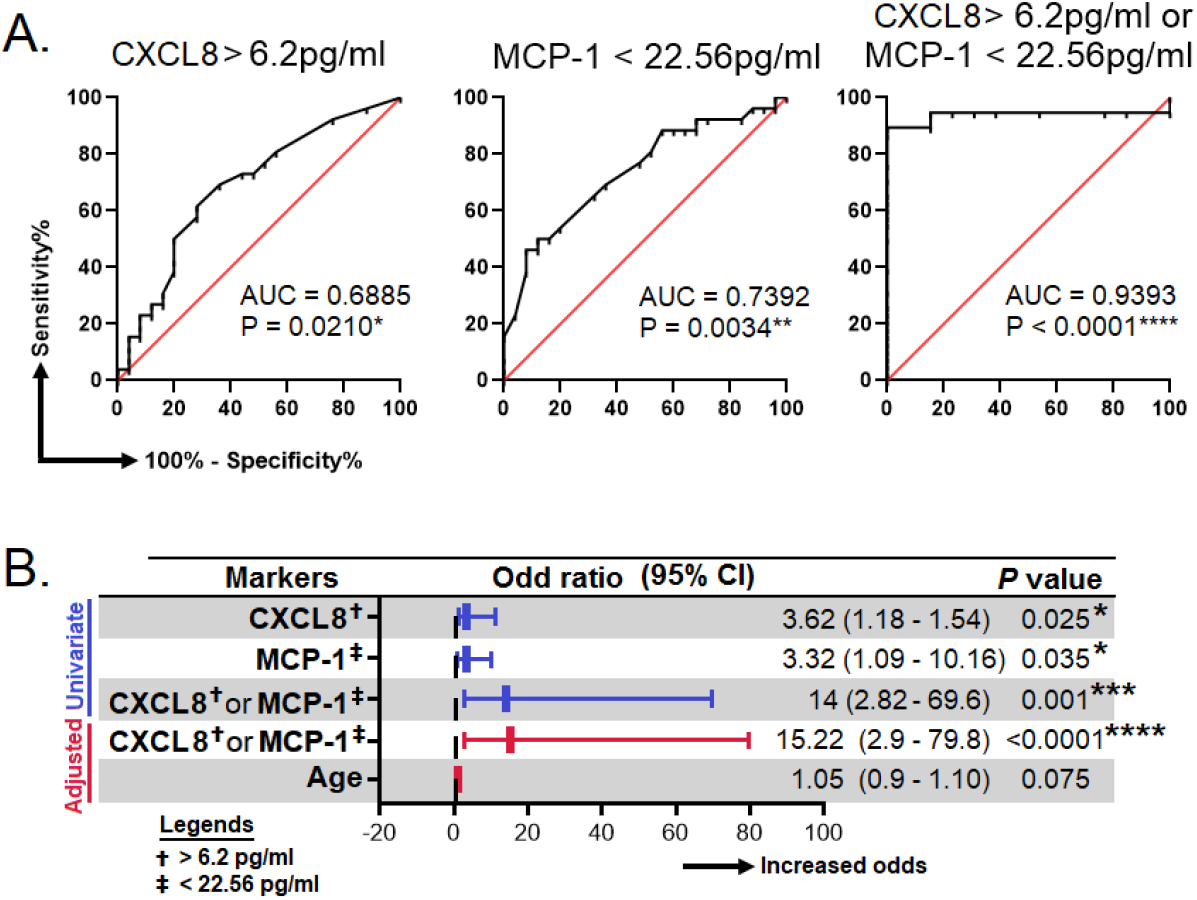
Efficacy of surrogate biomarkers in predicting LTBI. **A)** Receiver operating characteristic curves for the prediction of LTBI by using IL-8, MCP-1 and, either IL-8 or MCP-1. **B)** Association of IL-8, MCP-1 and, either IL-8 or MCP-1 with the risk of LTBI. These analyses were done between HC (true negative) and HCC/IGRA+ve (true LTBI). AUC, area under curve; CI, confident interval; IL, interleukin; MCP-1, monocyte chemoattractant protein-1. *, **, *** and, **** represent *p*<0.05, <0.01, <0.001 and <0.0001, respectively.

A binary regression analysis was performed to examine further the relationship between CXCL8 and MCP-1 and the likelihood of developing LTBI. We found that a combination of both CXCL8 (>6.2 pg/ml) and MCP-1 (<22.56 pg/ml) was associated with increased risk of LTBI by 14-fold (95% CI=2.82-69.6); *p*=0.001. Considering the levels of cytokines, which may change as age increases, we performed a multivariate binary regression analysis adjusting for age. The results showed that the CXCL8 (>6.2 pg/ml) and MCP-1 (<22.56 pg/ml) remained significant even with the change of age, where the odds was 15.22 (95% CI=2.9-79-8); *p*<0.0001 (*Figure 4B*).

## 4 DISCUSSION

LTBI has become a severe public health concern, not only because of the number of people that have already become latently infected with MTB but because of the risk of reactivation. >80% of the active TB cases reported in the US are due to reactivation, which can be prevented if effective screening tools are available to initiate timely treatment.^17^ An ideal biomarker needs to identify individuals with LTBI without needing *ex vivo* antigenic stimulation. This is because stimulation with MTB antigens such as PPD, CFP-10, and ESAT-6 would either have a strong cross-reactivity with BCG-vaccinated individuals or with someone with a history of MTB and hence skewing towards false-positive results. Furthermore, *ex vivo* stimulation usually does not work well in immunosuppressed or HIV-infected individuals.^14^

Here, we examined a fleet of cytokines for their potential to predict LTBI without *ex vivo* antigen stimulation. We identified two potential surrogate markers, CXCL8 and MCP-1, that could be combined to achieve an overall efficacy of ∼90%. As these markers could distinguish LTBI from HC without *ex vivo* antigen stimulation, using these markers may be cost-effective and less laborious, especially in resource-limited settings.^18^ An important mechanism in TB pathogenesis is granuloma formation, a cellular response in which macrophages accumulate locally in the lungs along with other leukocytes to “wall off” MTB from disseminating to host tissues.^19^ This is a complex process where multiple chemokines, especially CXCL8 and MCP-1 (or CCL-2), regulate leukocyte influx to the TB-infected sites.^20,21^ However, MTB is a highly successful pathogen that has evolved several strategies to evade host immunosurveillance to ensure its persistence in the host. The early secreted antigenic target 6 kDa (ESAT-6) is a virulence factor in MTB that significantly influences disease pathogenesis. ESAT-6 inhibits functional antigen-presenting cell responses by reducing IL-12 production by macrophages^22^ via their lysis,^23,24^ destabilizing phagolysosome to allow MTB to escape phagosome,^25^ and promoting their intracellular dissemination.^24,26^ Dissemination of ESAT-6 within macrophage cytosol could block the interaction between MyD88 and IRAK4 to prevent NF-κB activation from causing the attrition of IL-12, IL-6, IFN-γ, and TNF-α.^27,28^

Studies have shown that exposure to THP-1 with MTB and serum from active TB patients was associated with elevated CXCL8 and MCP-1 levels.^29^ However, we showed that individuals with LTBI had increased plasma CXCL8 but decreased MCP-1 levels. Such difference suggests that the MTB in HHCs might have altered the host immune responses to establish latency. Our observation that both IFN-γ and TNF-α were lower in LTBI individuals as compared to HCs justifies our finding. Others showed that mycobacteria-specific antigen-induced CXCL8 reduced with preventive treatment, offering hints for investigating prognostic biomarkers to assess performance. However, the CXCL8 levels were higher in both IGRA positive and negative groups than HCs in the unstimulated plasma, suggesting that CXCL8 might be secreted due to an ongoing MTB infection. Further, our binary regression analysis showed that the combination of CXCL8 and MCP-1 was associated with an increased risk of LTBI by 14-fold. Also, HHC/IGRA-ve individuals, albeit IGRA was negative, their cytokines profile was akin to HHC/IGRA+ve individuals indicating that they may have been in contact with MTB or LTBI. Though this shows the potential role of CXCL8 and MCP-1 in LTBI, our studies warranted large-scale epidemiological studies involving diverse groups of people and cohorts for the utility of CXCL8 and MCP-1 as a biomarker for LTBI.

## 5 CONCLUSIONS

In conclusion, our results identified CXCL8 and MCP-1 that could help identify LTBI cases. The combination of both CXCL8 and MCP-1 increased the risk of LTBI among HHCs 14-fold. Together, it could be construed that CXCL8 and MCP-1 could serve as surrogate biomarkers of LTB disease. The role of CXCL8 and MCP-1 as surrogate biomarkers warrant further validation for the possible detection of individuals with LTBI in the general population.

## Data Availability

All relevant data are within the paper and its Supporting information files.

## AUTHOR CONTRIBUTIONS

All authors have substantially participated in the preparation and agree to be accountable for all aspects of the work related to the manuscript. S.T.S., S.R., E.M.S., Y.K.Y., and V.V designed the study. B.A., A.F., L.M., M.R., A.M., K.S.J., V.K., A.A., and S.T.S., coordinated patient recruitment and sample collections. B.A., V.K., K.S.J., M.R., and A.A., developed the laboratory works and coordinated the development of the study. Y.K.Y., E.M.S., V.V., M.L., M.K., M.P.S., N.G., S.N.B., Y.Z., H.Y.T., R.V., and S.G performed data analysis. Y.K.Y., V.V., and E.M.S. did the biostatistical analysis. E.M.S., M.L., S.R., V.V., R.V., and M.L. co-wrote the manuscript. All authors reviewed and revised the manuscript critically.

## ACKNOWLEDGMENTS

The authors are grateful to all the participants, paraclinical, and laboratory staff of the National Tuberculosis Elimination Programme, Tamil Nadu, Chennai, and B. Kavitha and D. K. Mageshwari from the Tamil Nadu Public Health Laboratory, Directorate of Public Health and Preventive Medicine, Chennai, for assistance with patient recruitment, specimen collection, and cooperation.

